# White Matter microstructure effect in ADHD: a two-sample mendelian randomization study

**DOI:** 10.1101/2022.12.05.22282970

**Authors:** Maria Eduarda de Araujo Tavares, Marina Xavier Carpena, Eduardo Schneider Vitola, Cibele Edom Bandeira, Renata Basso Cupertino, Eduarda Colbeich, Pamela Ferreira da Cunha, Diego Luiz Rovaris, Eugenio Horacio Grevet, Bruna Santos da Silva, Claiton Henrique Dotto Bau

## Abstract

**Introduction:** Genome Wide Association Studies (GWAS) revealed the highly polygenic architecture of Attention-Deficit/Hyperactivity Disorder (ADHD) and highlighted the contribution of common variants related to brain development and function. In parallel, several imaging studies attempted to discover disorder-related brain structures, with some significant findings concerning white matter. Two-sample mendelian randomization (2SMR) is a powerful tool to evaluate causality between two phenotypes using summary statistics data. We aimed to investigate a possible causal relationship between white matter genetically predicted variation and ADHD diagnosis through 2SMR.

**Methods:** A unidirectional two-sample MR analysis was performed based on summary statistics of GWAS between 22 different white matter (WM) mean fractional anisotropy measures and ADHD. We used 4 different MR approaches, considering IVW random effects as the main analysis, followed by several sensitivity analyses. Linkage Disequilibrium Score Regression (LDSC) was evaluated in the same set of samples to corroborate the direction of associations.

**Results and Discussion:** Our most consistent finding across MR and LDSC approach, following the sensitivity analyses, indicate that the decreased WM microstructure integrity of the fornix stria terminalis (FXST_ivw_ beta:-0.266 SE:0.083 p_FDR:_ 0.021) genetic liability has a causal influence on ADHD diagnosis. The FXST is formed by connection fibers inside the limbic system, which is crucial to emotional processing, learning, and memory, functions usually impaired in ADHD. Therefore, this study increases knowledge concerning ADHD neurobiology and provides novel evidence of the causal effect of WM integrity in the limbic system, which could contribute to the advances in additional diagnostic tools as well as pharmacological brain structure targets.

## Introduction

Attention-Deficit Hyperactivity/Disorder (ADHD) is a neurodevelopmental disorder characterized by high levels of inattention, hyperactivity, and impulsivity, broadly impairing different aspects of life (American Psychiatric Association (APA), 2014). Although ADHD has one of the highest heritabilities (∼74%) across psychiatric disorders (Faraone & Larsson, 2019), specific pathophysiological mechanisms still need to be clarified. GWAS have revealed the highly polygenic architecture of ADHD, including the contribution of common variants related to brain development and function (Demontis et al. 2019. Rovira et al. 2020, Demontis et al., 2022).

Imaging studies with a broad range of methodologies attempt to discover disorder-related brain structures and explore the genetic variants associated with these phenotypes (Hoogman et al., 2017, 2019). Magnetic Resonance Imaging (MRI) scans offer a variety of structural brain analyses, including the Diffusion Tensor Imaging (DTI) technique. Through DTI, it is possible to observe the displacement of water molecules in the White Matter (WM) brain component and obtain derivative measures such as Fractional Anisotropy (FA), a value representing the orientation of water diffusion used to assess the integrity and direction of WM tracts (Le Bihan et al., 2001). WM connects the gray matter structures comprising cell membranes, glia, myelin, dendrites, and axon fibers (Adisetiyo, 2018), and it has a polygenic architecture supported by GWAS with common genetic variants explaining, on average, 41% of the population variation (B. Zhao et al., 2021).

Neuroimaging studies exploring DTI measures in patients with ADHD compared to typical development controls discovered some WM structures associated with the disorder, where the most robust findings are specific regions in the corpus callosum (Albajara Sáenz et al., 2019; Y. Zhao et al., 2022). Besides, DTI measures of the external capsule are highly correlated with ADHD pathophysiology. (B. Zhao et al., 2021). As DTI studies usually rely on small samples and different image acquisition parameters, processing, and analysis (Pereira-Sanchez & Castellanos, 2021), they have been susceptible to conflicting results (Y. Zhao et al., 2022). Since most studies evaluating imaging features in ADHD have an observational cross-sectional design (Firouzabadi et al., 2022), they are more susceptible to random and non-random bias including confounding bias which may limit the ability to infer causality. Two-sample mendelian randomization (2SMR) analysis emerges as a promising way to overcome these limitations. In 2SMR, two different summary datasets from independent samples are used to calculate the SNP-exposure and the SNP-outcome effects, which are used as instrumental variables (IV) to estimate the causal relationship of exposure on the outcome (to further insights into MR methodologies, see Supplementary Methods). Causal inferences between two traits can be made even if they are not measured in the same set of samples using only summary data, enabling harnessing the statistical power of pre-existing large GWAS analyses (Hemani et al., 2018; Pierce & Burgess, 2013). MR analyses require three major assumptions to have a robust causality inference. These include: relevance - the instrumental variables (SNPs) have to be associated with the exposure; exchangeability - the instrumental variables affect the outcome only through the exposure; exclusion restriction - the absence of a confounder affecting the exposure-outcome relationship (VanderWeele et al., 2014).

The violation of these assumptions can limit the power and reliability of the MR results. Therefore, it is a huge challenge to investigate highly polygenic and pleiotropic phenotypes such as brain connectivity measures and mental disorders diagnoses due to the difficulty in addressing these premises. In this sense, new methods have been applied to assess the degree of horizontal pleiotropy between traits, e.g. linkage disequilibrium score regression (LDSC - Bulik-Sullivan et al., 2015), helping to evaluate the degree and direction of genetic correlation, besides several sensitivity analyses (Burgess et al., 2017).

This study relied on the reported influence of white matter microstructure integrity in ADHD etiology, the difficulty of gathering DTI studies results, the possibility raised by new methods to combine different GWAS information, and the availability of large samples with ADHD and FA data. In this scenario, we tested for a possible causal relationship between brain white matter microstructure and ADHD by evaluating causality and pleiotropy, throughout 2SMR and LDSC.

## Methods

### Samples

We used two summary level GWAS data: 1) the first provided FA WM microstructure measures of 21 tracts and one global measure (anterior corona radiata - ACR; anterior limb of internal capsule - ALIC; Average Fractional Anisotropy; body of corpus callosum- BCC; cingulum (cingulate gyrus) - CGC; cingulum (hippocampus) - CGH; corticospinal tract - CST; external capsule - EC; fornix (column and body of fornix) - FX; fornix-stria terminalis - FXST; genu of corpus callosum - GCC; inferior fronto-occipital fasciculus - IFO; posterior corona radiata - PCR; posterior limb of internal capsule - PLIC; posterior thalamic radiation - PTR; retrolenticular part of internal capsule - RLIC; splenium of corpus callosum - SCC; superior corona radiata - SCR; superior fronto-occipital fasciculus - SFO; superior longitudinal fasciculus - SLF; sagittal stratum - SS; Uncinate Fasciculus - UNC) of 33,292 subjects evaluated in the UK Biobank (Zhao et al. 2021); 2) the second was the clinically diagnosed ADHD versus controls, including 53,293 subjects conducted by Demontis and colleagues in 2019. Both are the largest GWAS of WM and ADHD with summary statistics available until the assembly of this paper. Summary GWAS data are available at http://doi.org/10.5281/zenodo.4549730.129; and https://www.med.unc.edu/pgc/download-results/. The samples did not have substantial overlap and were mostly of European descent.

### 2SMR

We analyzed the causal relationship between 22 mean FA measures assessed through WM GWAS (Zhao et al., 2021) and ADHD diagnosis (Demontis et al., 2019) using four different MR approaches (1. MR-Egger, 2. Weighted Median, 3. IVW - Inverse Variance Weighted, and 4. GSMR). We considered random effects IVW as the main analysis and concordant results between approaches as a robustness indicator. Each approach has a different way of dealing with the MR assumptions described above and detailed in the Supplementary Methods. To select our instrumental variables (IV) we clumped the WM summary statistics selecting SNPs reaching genomic significance (p < 5E-08) with a 1000 kb window and a r² = 0.01 for each tract selected as exposure. Then, we harmonized the exposure beta and standard error coefficients with those calculated for the SNPs associated with the outcome (ADHD) to perform the MR. Ambiguous and palindromic SNPs were excluded. False Discovery Rate (FDR) was applied to multiple test correction (Benjamini & Hochberg, 1995).

### Sensitivity Analyses

We estimated the validity of the IV included in the 2SMR through F-statistics and I² to test the violation of no measurement error (NOME) on the SNP-exposure association. After that, we conducted several sensitivity analyses to assess if any MR assumptions were violated, including Cochran’s Q Statistics, MR Egger regression, Steiger directionality test, MR-PRESSO, MR-RAPS, Leave-one-out analysis, Funnel Plots, and HEIDI-test in all tracts included in the 2SMR analyses. Statistical power was estimated through the online tool <u>mRnd: Power calculations for Mendelian Randomization</u> (shiny.cnsgenomics.com), further described in (Brion et al., 2013).

### LDSC

LDSC (Bulik-Sullivan et al., 2015) was used to assess the degree and direction of genetic correlation between brain white matter (Zhao et al. 2021) and ADHD GWAS (Demontis et al., 2019), in addition to the inter-correlations between each tract. FDR was applied to multiple test correction (Benjamini & Hochberg, 1995).

We used the TwoSampleMR R package (https://github.com/MRCIEU/TwoSampleMR - DOI: https://doi.org/10.7554/eLife.34408). and the Complex-Traits Genetics Virtual Lab (https://www.biorxiv.org/content/10.1101/518027v4 - DOI: https://doi.org/10.1101/518027) default parameters to perform all analyses. We followed the STROBE-MR (Skrivankova et al., 2021) and complementary guidelines (Burgess et al., 2019; Skrivankova et al., 2021) to assemble and present our results.

## Results

Our analyses flow is summarized in Figure 1.

**Figure 1.**
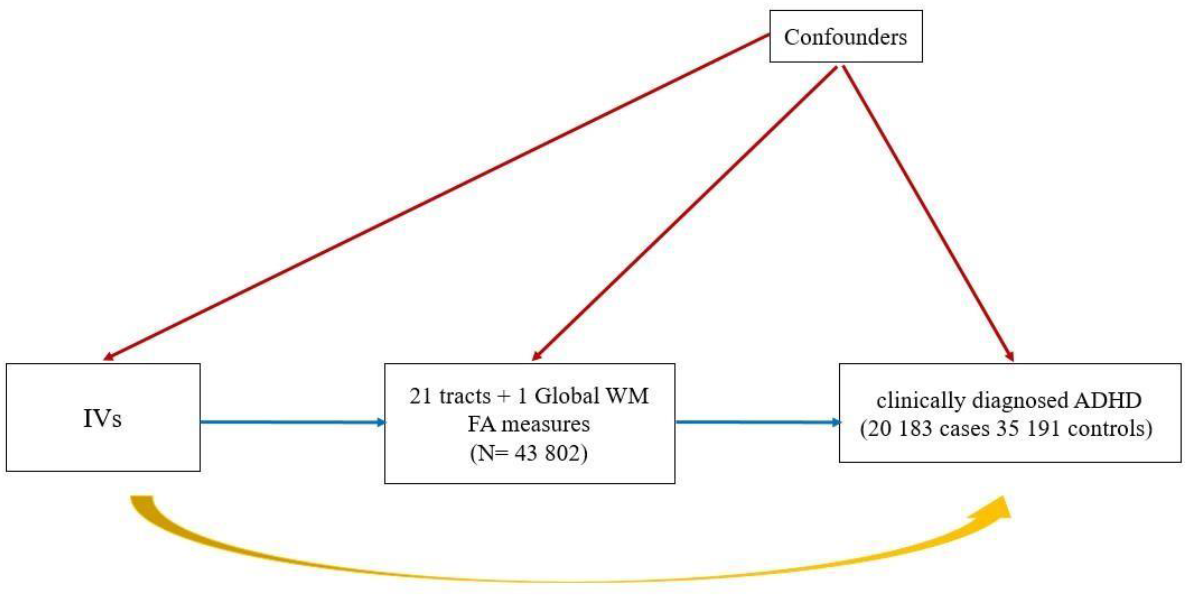
The three main assumptions of unidirectional two-sample MR are shown in blue relevance - the instrumental variables (SNPs) have to be associated with the exposure); yellow (exchangeability - the instrumental variables affect the outcome only through the exposure); and red (exclusion restriction - the absence of a confounder affecting the exposure-outcome relationship). The relevance assumption was addressed by the MR analyses performed, including as IVs only independent SNPs highly associated (5e10^−8^) with the FA measures with validity confirmed by I² measure, F-statistic, heterogeneity tests, leave-one-out, and MR-RAPS analyses. Exchangeability was assessed through the sensitivity analyses evaluating pleiotropy (MR-Egger regression, HEIDI-test, MR-PRESSO), and directionality (Steiger test). Exclusion restriction was estimated through funnel plots.

### 2SMR

We included the 22 WM tracts FA measures as exposures in different models of 2SMR analyses to estimate the causal effect of each WM on ADHD. We excluded CST in this analysis as this structure did not have sufficient associated SNPs to perform the models selected. Our main analyses (IVW) show that CGH (beta: −0.379, SE: 0.122) and FXST (beta: −0.266 SE:0.083) have significant associations with ADHD after FDR correction (Table 1). In addition, another four tracts (ACR, ALIC, BCC, GCC) were significant in at least 2 MR methods. Results for all tracts and MR approaches are compiled in Table 1.

**Table 1.**
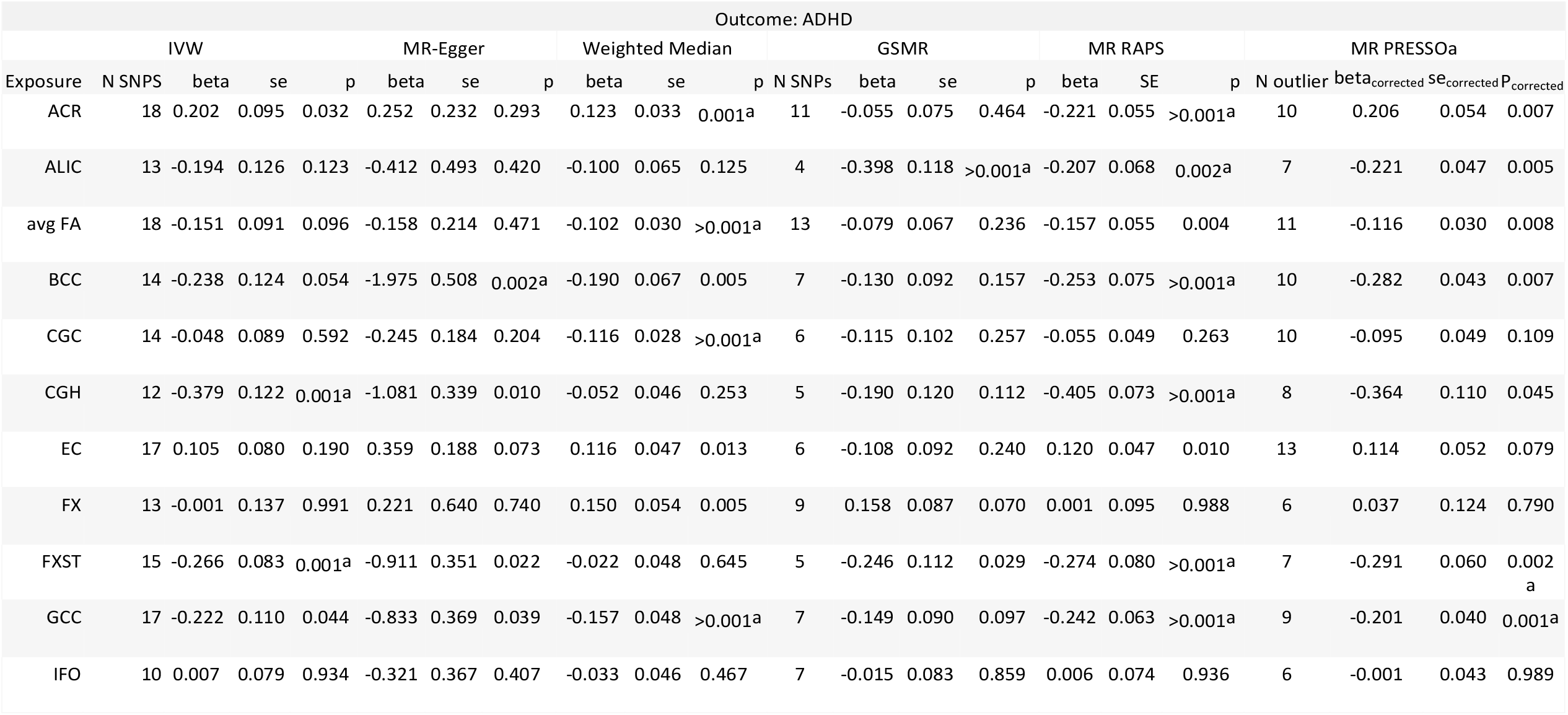
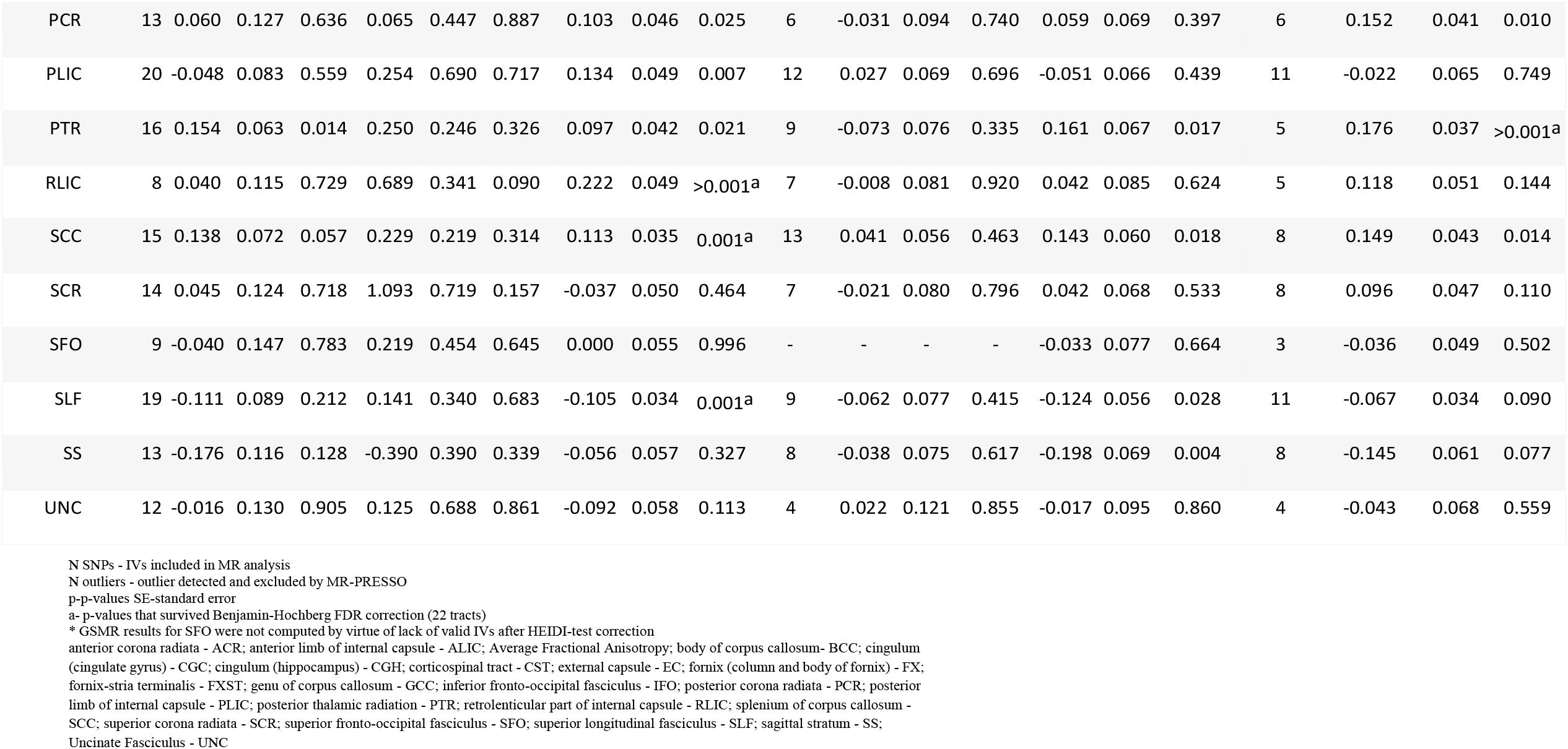
2SMR analyses results

### Sensitivity Analyses

The mean F-statistics calculated for all tracts are considerably high with values >30 represent good indications of the instrumental variable’s viability. The tracts IVs were closely between the range of 0.6 > I²_GX_ > 0.9 proposed by (Bowden et al., 2015) (Table 2). MR-Egger intercept was relatively close to zero, with indication of pleiotropy only in BCC. Heterogeneity tests (MR-Egger and IVW) were significant for all tracts, and forest plots also indicated heterogeneity across IVs, with probable violation of the MR assumptions. The leave-one-out analysis did not indicate any specific variant carrying the effect found in the MR analysis of CGH and FSXT. Funnel plots evidence overdispersion of the data for ALIC, BCC, and GCC. Steiger sensitivity test also corroborates the exposure upstream of the outcome for all structures, indicating no reverse causation. SFO was excluded from GSMR in the absence of valid IVS after HEIDI-test. Concerning our main analysis results, FXST tract has pleiotropic variants indicated through MR-PRESSO global test of pleiotropy, but the analysis remained significant after the outlier correction, which is corroborated by the MR-RAPS results. In the same sense, the tract remained significant in GSMR (p<0.05) after the HEIDI-test of pleiotropy. CGH tract did not survived GSMR correction for pleiotropy and remained significant in MR-PRESSO and MR-RAPS models. The other tracts have heterogenous results, but all were significant for MR-RAPS and at least one of the other methods. ALIC was the only significant (p FDR) for GSMR. The power of MR analysis to establish causality was 100%. All results are compiled in Table 2.

**Table 2.**
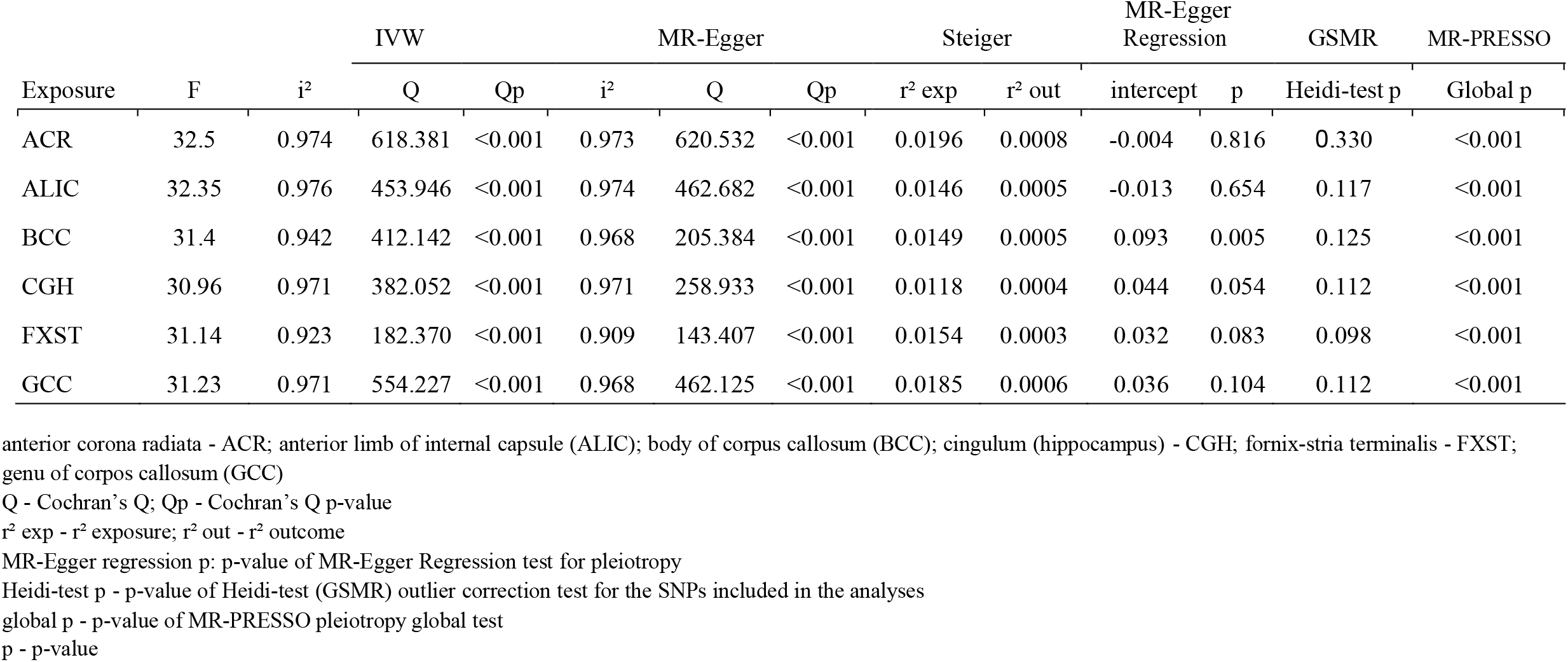
Sensitivity analyses results for ACR, ALIC, BCC, CGH, FXST and GCC (tracts associated by at least 2 MR analyses), other results are compiled in the Supplementary Material.

| Exposure | F | i<sup>2</sup> | IVW |  | MR-Egger |  |  | Steiger |  | MR-Egger Regression |  | GSMR | MR-PRESSO |
| --- | --- | --- | --- | --- | --- | --- | --- | --- | --- | --- | --- | --- | --- |
|  |  |  | Q | Qp | i<sup>2</sup> | Q | Qp | r<sup>2</sup> exp | r<sup>2</sup> out | intercept | p | Heidi-test p | Global p |
| ACR | 32.5 | 0.974 | 618.381 | <0.001 | 0.973 | 620.532 | <0.001 | 0.0196 | 0.0008 | -0.004 | 0.816 | 0.330 | <0.001 |
| ALIC | 32.35 | 0.976 | 453.946 | <0.001 | 0.974 | 462.682 | <0.001 | 0.0146 | 0.0005 | -0.013 | 0.654 | 0.117 | <0.001 |
| BCC | 31.4 | 0.942 | 412.142 | <0.001 | 0.968 | 205.384 | <0.001 | 0.0149 | 0.0005 | 0.093 | 0.005 | 0.125 | <0.001 |
| CGH | 30.96 | 0.971 | 382.052 | <0.001 | 0.971 | 258.933 | <0.001 | 0.0118 | 0.0004 | 0.044 | 0.054 | 0.112 | <0.001 |
| FXST | 31.14 | 0.923 | 182.370 | <0.001 | 0.909 | 143.407 | <0.001 | 0.0154 | 0.0003 | 0.032 | 0.083 | 0.098 | <0.001 |
| GCC | 31.23 | 0.971 | 554.227 | <0.001 | 0.968 | 462.125 | <0.001 | 0.0185 | 0.0006 | 0.036 | 0.104 | 0.112 | <0.001 |

anterior corona radiata - ACR; anterior limb of internal capsule (ALIC); body of corpus callosum (BCC); cingulum (hippocampus) - CGH; fornix-stria terminalis - FXST; genu of corpus callosum (GCC)
Q - Cochran's Q; Qp - Cochran's Q p-value
r<sup>2</sup> exp - r<sup>2</sup> exposure; r<sup>2</sup> out - r<sup>2</sup> outcome
MR-Egger regression p: p-value of MR-Egger Regression test for pleiotropy
Heidi-test p - p-value of Heidi-test (GSMR) outlier correction test for the SNPs included in the analyses
global p - p-value of MR-PRESSO pleiotropy global test
p - p-value

### LDSC

Amongst the 22 tracts evaluated, eight were negatively correlated (p<0.05) with ADHD, including the average FA global measure, GCC, CGH, ALIC, and FXST. None of the tracts survived the FDR correction (Table 3, Figure 2). The pairwise tract correlations are presented in Figure 3. Almost all white matter integrity measures present high positive genetic correlations (rG>0.6), as expected for different component proxies of the same biological structure. However, some correlations are weaker or even negative, exhibiting the highly complex and heterogeneous genetic compound of brain WM microstructure. CGH and FXST were weakly correlated, demonstrating the independence of MR results (Figure 3).

**Table 3.** LDSC results for WM tracts and ADHD

| WM Structure | LDSC |  |  |  |  |  |
| --- | --- | --- | --- | --- | --- | --- |
| | $h^2$ SNP | $h^2$ SNP SE | rG | SE | p | pFDR |
| ACR | 0.360 | 0.03 | -0.08 | 0.044 | 0.080 | 0.195 |
| ALIC | 0.280 | 0.024 | -0.053 | 0.044 | 0.235 | 0.323 |
| avg FA | 0.320 | 0.030 | -0.106 | 0.043 | 0.014 | 0.077 |
| BCC | 0.310 | 0.032 | -0.025 | 0.044 | 0.567 | 0.656 |
| CGC | 0.360 | 0.030 | -0.016 | 0.041 | 0.7 | 0.733 |
| CGH | 0.270 | 0.022 | -0.060 | 0.043 | 0.165 | 0.259 |
| CST | 0.190 | 0.020 | -0.067 | 0.056 | 0.227 | 0.323 |
| EC | 0.310 | 0.030 | -0.068 | 0.045 | 0.127 | 0.233 |
| FX | 0.250 | 0.025 | 0.036 | 0.049 | 0.460 | 0.562 |
| FXST | 0.310 | 0.027 | -0.048 | 0.045 | 0.296 | 0.383 |
| GCC | 0.360 | 0.030 | -0.115 | 0.041 | 0.005 | 0.055 |
| IFO | 0.350 | 0.036 | 0.065 | 0.046 | 0.155 | 0.259 |
| PCR | 0.290 | 0.024 | -0.090 | 0.042 | 0.031 | 0.100 |
| PLIC | 0.260 | 0.026 | -0.074 | 0.047 | 0.113 | 0.226 |
| PTR | 0.270 | 0.026 | -0.115 | 0.041 | 0.005 | 0.055 |
| RLIC | 0.270 | 0.026 | -0.101 | 0.047 | 0.032 | 0.100 |
| SCC | 0.250 | 0.031 | -0.015 | 0.046 | 0.738 | 0.738 |
| SCR | 0.280 | 0.025 | -0.104 | 0.047 | 0.025 | 0.100 |
| SFO | 0.230 | 0.024 | -0.095 | 0.047 | 0.044 | 0.121 |
| SLF | 0.350 | 0.027 | -0.066 | 0.04 | 0.100 | 0.220 |
| SS | 0.270 | 0.029 | -0.114 | 0.043 | 0.008 | 0.058 |
| UNC | <u>0.240</u> | <u>0.026</u> | <u>-0.020</u> | <u>0.050</u> | <u>0.685</u> | <u>0.733</u> |

$h^2$ SNP - SNP heritability; $h^2$ SNP SE - standard error
rG - correlation r SE - standard error
p - p-value pFDR- False Discovery Rate corrected p-value

**Figure 2.**
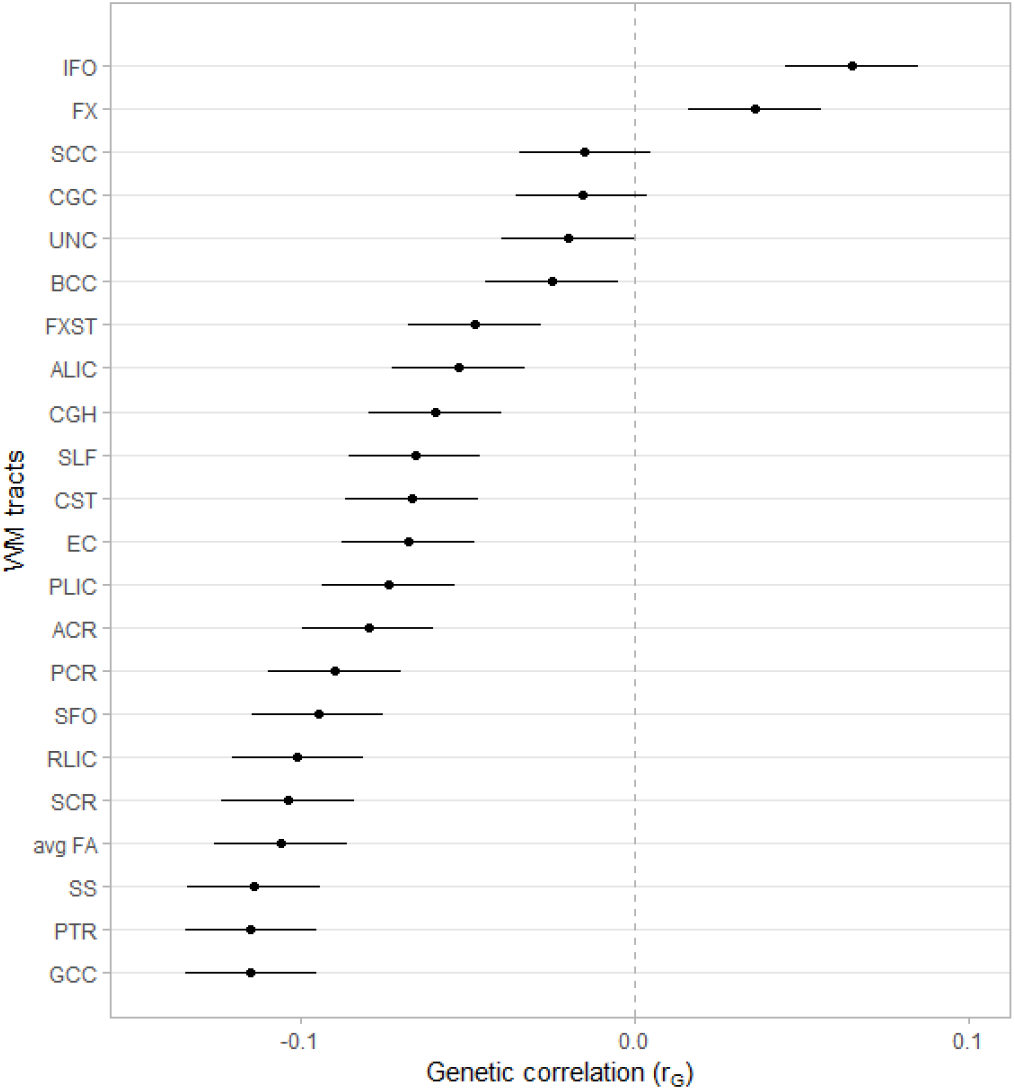
Genetic correlation between 22 FA measures (Zhao et al. 2021) and ADHD (Demontis et al. 2019) Correlations between ADHD and 22 White Matter fractional anisotropy measures anterior corona radiata - ACR; anterior limb of internal capsule - ALIC; Average Fractional Anisotropy; body of corpus callosum-BCC; cingulum (cingulate gyrus) - CGC; cingulum (hippocampus) - CGH; corticospinal tract - CST; external capsule - EC; fornix (column and body of fornix) - FX; fornix-stria terminalis - FXST; genu of corpus callosum - GCC; inferior fronto-occipital fasciculus - IFO; posterior corona radiata - PCR; posterior limb of internal capsule - PLIC; posterior thalamic radiation - PTR; retrolenticular part of internal capsule - RLIC; splenium of corpus callosum - SCC; superior corona radiata - SCR; superior fronto-occipital fasciculus - SFO; superior longitudinal fasciculus - SLF; sagittal stratum - SS; Uncinate Fasciculus - UNC

**Figure 4.**
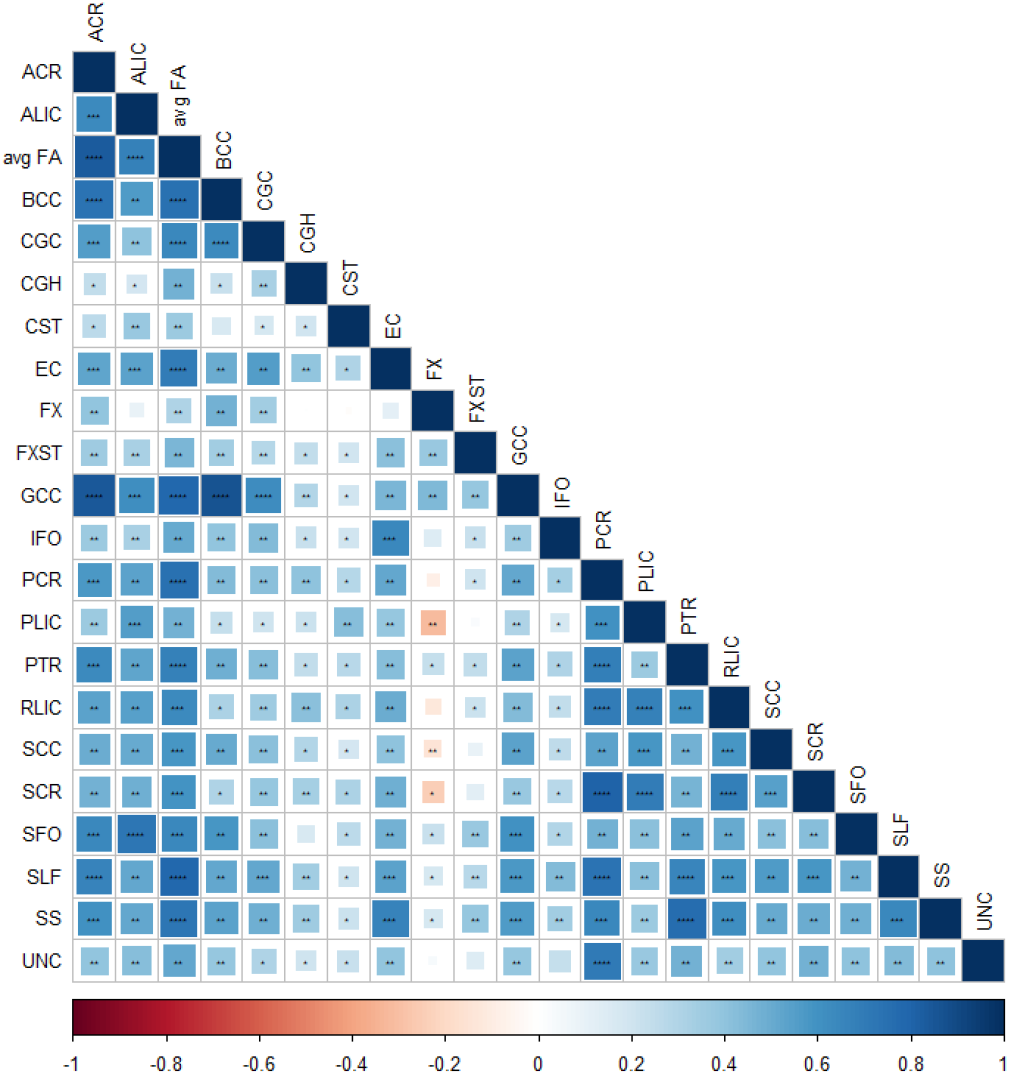
Pairwise correlation plot between WM tracts fractional anisotropy measures evaluated in 33,292 individuals in the UKBiobank (Zhao et al. 2021) Pairwise correlations between 22 White Matter fractional anisotropy measures * - 0.002 ** - 5×10^−10^ *** - 5×10^−50^ **** - 5×10^−100^ anterior corona radiata - ACR; anterior limb of internal capsule - ALIC; Average Fractional Anisotropy; body of corpus callosum- BCC; cingulum (cingulate gyrus) - CGC; cingulum (hippocampus) - CGH; corticospinal tract - CST; external capsule - EC; fornix (column and body of fornix) - FX; fornix-stria terminalis - FXST; genu of corpus callosum - GCC; inferior fronto-occipital fasciculus - IFO; posterior corona radiata - PCR; posterior limb of internal capsule - PLIC; posterior thalamic radiation - PTR; retrolenticular part of internal capsule - RLIC; splenium of corpus callosum - SCC; superior corona radiata - SCR; superior fronto-occipital fasciculus - SFO; superior longitudinal fasciculus - SLF; sagittal stratum - SS; Uncinate Fasciculus - UNC

## Discussion

Our study is the first to evaluate the causal relationship and genetic correlation between a wide range of white matter microstructure features and ADHD. Among the 22 WM FA measures analyzed, the FXST presented the most consistent evidence of a causal effect on ADHD, expressed as a diminished WM integrity of this tract related to an increased ADHD risk. The robustness of this association is reinforced by the cross-method concordance and validation through sensitivity and posthoc analyses. These findings on causal effects in ADHD support previous evidence pointing the white matter microstructure of the limbic circuitry in both relevant dimensional traits involved in emotional regulation and the susceptibility to psychiatric disorders (Kebets et al., 2021).

We performed a unidirectional MR analysis assuming that, from a temporal perspective, genetically driven WM microstructure development occurs before ADHD appearance. We found evidence of MR assumption violation in all tracts through Cochrane’s Q heterogeneity test, which can be a source of bias in the results. Nevertheless, after a range of sensitivity analyses accounting for pleiotropy and IVs outliers (MR-PRESSO, MR-RAPS, and HEIDI-test) the results found for FXST remained significant across most methods (p<0.05). The CGH was not associated through GSMR after the HEIDI test of pleiotropy, which we consider as a probable bias indicative. We also present the genu of corpus callosum (GCC) and the anterior limb of internal capsule (ALIC) WM regions as secondary promising results, since they were consistent across methods that better accounts for MR violations (such as horizontal pleiotropy), even not being associated through our main analysis.

FXST, CGH, and GCC are all components of the Papez circuit formed by the limbic white matter tracts (Koshiyama et al., 2020; Shah et al., 2012). FXST and CGH are part of concentric rings surrounding the thalamus, separated by the corpus callosum, connecting the temporal pole to other limbic centers (Pascalau et al., 2018), while ALIC is part of the temporal stem which connects the frontal and temporal lobes (Peltier et al., 2010). The limbic system modulates memory, emotions, and behavior, and some of its regions, such as the amygdala, accumbens, and hippocampus have been found to be smaller in ADHD subjects than controls (Hoogman et al., 2017). Besides, white matter microstructure alterations of the limbic circuitry components found to be associated with ADHD in this study have been related to other brain disorders, as Alzheimer’s (Dalboni da Rocha et al., 2020), schizophrenia, and bipolar disorder (Koshiyama et al., 2020).

Genetic correlations can indicate pleiotropy or causal association loci in two traits, a combination of vertical (exists a causal relationship between two phenotypes where one leads to an increased risk of the other), and horizontal pleiotropy (the genetic variant contributes directly or indirectly to the risk of both phenotypes) (van Rheenen et al., 2019). The LDSC results suggest some overlap between WM tracts and ADHD genetics, with most tracts FA being negatively correlated with the disorder. This is expected and concordant with the literature, and our 2SMR findings, although some studies have conflicting results (Albajara Sáenz et al., 2019; Y. Zhao et al., 2022). We also evaluated the degree and direction of genetic correlations across tracts, certifying that our results were not signals of associations within the structures.

Our study must be seen considering some strengths and limitations. The summary statistics used were the largest ones publicized so far for both phenotypes, with minimum sample overlap, alongside an MR power close to 100%. We evaluated reverse causation and other violations of MR assumptions that were discarded or corrected by comprehensive sensitivity analyses. The comparability with the previous study conducted by Zhao et al., (2021) is impaired because we focused on FA measures while they used 171 DTI centered measures, including other parameters beyond FA, divided into 5 principal components. This could explain the differences in the WM structures genetically correlated with ADHD observed between the studies. Another fact to consider while interpreting the results is that all genetic variants included in the analyses were based on associations from studies evaluating European descent individuals, therefore, our results cannot be extrapolated to other ethnicities.

In conclusion, the use of two-sample mendelian randomization with several sensitivity analyses enhances our ability to infer the causality of WM microstructure variation on ADHD. The concordance between MR models and LDSC based on complex associations between multifactorial and pleiotropic traits strongly indicates the robustness of the results. If replicated, the decreased WM integrity in the FXST region could act as an ADHD endophenotype and contribute to the future landscape of accessory diagnostic tools

## Data Availability

All data produced in the present study are available upon reasonable request to the authors

